# Exploratory Validation of a Survey Instrument Based on the Theory of Planned Behavior to Assess Vaping Attitude and Perceptions

**DOI:** 10.1101/2024.09.04.24313091

**Authors:** Muna Barakat, Roa’a Abuarab, Baraah Alkharabsheh, Nada Budair, Mais Fareed, Rahma Kharabsheh, Dana Oshroq Laban, Malik Sallam

## Abstract

The prevalence of vaping (e-cigarette use) surged among young individuals, especially university students, highlighting the necessity to explore the factors influencing this behavior. This study aimed to develop and validate the Vaping Attitude and Perceptions Scale (VAPeS), based on the Theory of Planned Behavior (TPB). A cross-sectional design was employed using a self-administered electronic questionnaire to collect data from university students in Jordan. The questionnaire assessed demographic variables and key constructs of the VAPeS: Social Influence, Perceived Harms, Pleasure Derived from Vaping, Behavioral Influences, and Economic Factors and Self-Efficacy. The survey engaged 671 university students, with 201 (30.0%) reporting the use of tobacco products. Among these tobacco users, a majority (n=126, 62.7%) reported the current use of e-cigarettes. The exploratory factor analysis (EFA) using Principal Component Analysis (PCA) revealed a five-factor solution explaining 64.5% of the variance. These factors—Social Influence, Perceived Harms, Vaping Pleasure, Behavioral Influences, and Economic and Self-Efficacy—demonstrated high internal consistency. Regression analysis showed that Social Influence, Behavioral Influences, and sex were significant predictors of vaping behavior, with standardized coefficients of 0.494, 0.206, and −0.154, respectively (*P*<.001, *P*=.008, and *P*=.018). The VAPeS displayed satisfactory psychometric properties, making it a valuable tool for investigating vaping behaviors among the youth. Factors such as Social Influence and Behavioral Influences were significant factors in shaping vaping behaviors among the participants. These insights emphasize the need to consider these dimensions in public health initiatives aimed at managing the rising vaping prevalence among university students. The VAPeS instrument can be utilized for better delineation of vaping behaviors among the younger demographic. Subsequently, the insights gained through VAPeS-based research can help devise targeted educational campaigns aimed at addressing the perceived social acceptability and the supposed safety of vaping.

## Introduction

Tobacco consumption is widely recognized as an important health hazard, contributing to a wide range of chronic diseases and fatal illnesses (e.g., lung cancer, chronic obstructive pulmonary disease, coronary artery disease, etc.) [1]. Nevertheless, the prevalence of tobacco use remains high in various countries, including Jordan, which is ranked among the top countries for tobacco consumption among males worldwide [2,3]. The complexity of this challenge needs to be addressed via multiple approaches and the behavioral aspects is of particular importance considering the central role of the behavioral factors in determining tobacco consumption [4,5].

Vaping (e-cigarette use) is a relatively new appealing form of tobacco use and it represents an additional challenge for the efforts aiming to reduce tobacco consumption and its associated harm [6–8]. In university students and particularly among students in health schools, the perception of vaping as a less harmful alternative to traditional smoking or as a helpful method for smoking cessation might influence their attitudes and behaviors towards this form of smoking, despite the emerging evidence of vaping health risks [9–12].

The Theory of Planned Behavior (TPB) is a well-established psychological framework that can be valuable in understanding the determinants of health behavior in various contexts [13–17]. This is particularly relevant upon studying a group like university students, including those studying in health schools with such students characterized by being well-informed about the health risks associated with smoking [18–26]. However, university students especially those studying in health schools, are often challenged by high levels of stress and anxiety related to academic pressure, which might influence their smoking behavior [27–29].

In Jordan among other countries and regions worldwide, the cultural and social norms can play a significant role in determining the smoking behavior, including vaping [30,31]. Thus, the TPB can be a helpful framework to understand the underlying factors driving such behavior [17]. According to the TPB, behavior is driven by three constructs: attitudes (the individual positive or negative attitude of the behavior), subjective norms (the perceived social pressure to engage in the behavior), and the perceived behavioral control (perceived ease of performing the behavior) [13].

University students in general might develop a positive attitude towards smoking as a coping mechanism for stress and anxiety, despite their knowledge of the health hazards of smoking [32,33]. This situation is further complicated by subjective norms in Jordan with high prevalence of smoking including the e-cigarette use [31,34]. In such a context, the social acceptance of smoking can be expected among families and peers, which can influence the individual behavior regarding smoking [35]. Moreover, the availability and accessibility of tobacco products and e-cigarettes, besides the perceived inability to resist social pressures or to find alternative stress-coping mechanisms, can be critical factors in the decision-making process of the general public as well as among university students [35,36].

Previous studies highlighted the rising trend of vaping in Jordan among other Middle East countries [8,12,31,37,38]. Therefore, the aim of this study was to evaluate the determinants of vaping among university students in Jordan. This pilot study can help to establish a survey instrument tailored to understand the behavioral determinants of vaping among university students in the Arab countries where the prevalence of vaping is rising [8,31,37]. Subsequently, the developed survey instrument can be used for a wider scale studies that can form the basis for evidence-based efforts to address the problem of smoking and the emerging issue of vaping in Arab countries.

The current study aimed to address significant gaps in the existing literature on e-cigarette use in the Middle East as elaborated on by Al-Hamdani & Brett Hopkins [37]. In their recent commentary, Al-Hamdani & Brett Hopkins highlighted the lack of standardized measurement instruments to assess e-cigarette use in the Middle East [37]. By employing a comprehensive survey instrument based on the TPB, our study aimed to delineate a broad spectrum of vaping attitudes and perceptions among university students in the Middle East through the use of a validated survey instrument which would set a new standard for consistency and comparability in future research, filling critical knowledge gaps and informing public health strategies regionally.

## Materials and Methods

### Study design and ethics statement

The study was designed as a cross-sectional survey utilizing an electronic format for data collection. The study was approved by the Institutional Review Board (IRB) at the Faculty of Pharmacy, Applied Science Private University (reference number: 2024-PHA-2). Providing an electronic informed consent was required for participants to complete the survey successfully, and no identifying information was collected.

### Sampling methodology

To expedite data collection, a convenience sampling strategy was implemented. The initiation of the survey distribution was supervised by two Associate Professors, each affiliated with different academic institutions in Jordan—one from a public university and another from a private university to reach a wide spectrum of potential participants. Further distribution was carried out by the co-authors, who are medical students, utilizing Facebook and WhatsApp groups, which are popular communication channels among university students in Jordan. Data collection started on 7 July 2024 and concluded on 17 August 2024 and no incentives were offered to recruit potential participants.

### Sample size calculation

To determine the minimum sample size for the estimation of the prevalence of vaping and tobacco consumption among university students in this study, we employed the Epitools online tool for estimating a proportion with specified precision [39]. The pre-defined parameters for this calculation included an estimated true proportion of 0.5, a precision of ±0.05 with a confidence level of 95%, considering the population size of about 350,000 university students [40]. This calculation yielded a required sample size of 385 participants. For the exploratory factor analysis (EFA) validation of the survey instrument used to assess the attitude to vaping, the sample size was derived based on analytical requirements specific to factor analysis. With 23 items in our survey, we targeted a minimum of 115 participants (5 participants × 23 items) [41].

### Survey instrument development

The survey items were initially developed in English and underwent a rigorous translation and back-translation process to ensure linguistic and conceptual accuracy in Arabic by both the first author and senior author, who are experts in tobacco research and survey methodology, respectively, and both bilingual in Arabic and English. This process aimed to preserve the content validity across linguistic variations. The survey was designed to explore various constructs pertinent to vaping based on the TPB constructs, including attitudes towards vaping, perceived social norms, and perceived behavioral control [13–17]. Pilot testing of the novel survey instrument was conducted on six medical students with subsequent feedback and minor editing to improve the clarity and readability of a few survey items.

The survey which was hosted on Google forms started by an introductory section on the study objectives and upon securing informed consent, demographic data were collected including age, sex, parental education, household income, and family tobacco use history, among others.

The survey included direct queries about tobacco usage as follows. (1) “Currently, do you smoke any kind (cigarettes, hookah, e-cigarettes (vape))?” and (2) “Are you currently using e-cigarettes (vaping)?”, with options for “Yes” or “No”.

Afterward, responses to 23 TPB-based attitudinal and behavioral items were introduced and measured using a 5-point Likert scale, ranging from “strongly agree” to “strongly disagree”. The constructs measured in the survey comprised the following: (A) Perceived Harm with three items: (1) I believe that vaping is less harmful compared to cigarettes; (2) I believe that vaping is less harmful compared to Narghile; and (3) E-cigarettes (vaping) can help smokers quit smoking or cut down on smoking.

(B) Perceived Enjoyment with two items: (1) I find vaping enjoyable; and (2) I find vaping satisfying. (C) Perceived Health Risks with three items: (1) I believe that vaping is associated with health risks; (2) I believe that vaping can lead to nicotine addiction; and (3) I am aware of anti-smoking policies within my university.

(D) Perceived Cost with two items: (1) I believe that vaping is less costly compared to cigarettes; and (2) I believe that vaping is less costly compared to Narghile. (E) Family and Peer Influence with five items: (1) My vaping experience is influenced by the behavior in my family; (2) I feel pressured to smoke by my peers; (3) My vaping experience is influenced by the behavior of my friends; (4) My role models in life consume tobacco in any form; and (5) Vaping is socially accepted among my peers.

(F) Media Influence with two items: (1) The portrayal of vaping in movies and series influences how I perceive it; and (2) The social media portrayal influence how I perceive vaping. (G) Confidence in Ability to Quit with two items: (1) I feel confident in my ability to resist vaping in social situations; and (2) I feel confident in my ability to quit vaping altogether. (H) Need to Cope with Stress with three items: (1) Academic stress affects my vaping habits; (2) I believe that vaping helps in social acceptance; and (3) I believe that vaping helps in stress reduction.

(I) Easy Access to Vaping with a single item: (1) I can easily obtain e-cigarettes.

### Statistical analysis

The collected data from the survey was subjected to EFA employing principal component analysis (PCA) as the extraction method to uncover latent structures within the data. To accommodate the anticipated correlations among factors, an Oblimin rotation was applied. To evaluate the internal consistency of the scales derived from EFA, the Cronbach’s α was calculated for each inferred construct. Each extracted construct was quantified by averaging the scores of the items within it, with responses ranging from “strongly agree” (scored as 5) to “strongly disagree” (scored as 1). For the final VAPeS score as averaged based on the average score for the 23 items, categorization into two categories was conducted as follows: (1) Agreement category with a VAPeS score range of 3.68–5.00; and (2) Disagreement/neutral category with a VAPeS score range of 1.00–3.67.

Associations between categorical variables were assessed using the Chi-square (χ^2^) test. For associations involving categorical and scale variables, normality was first checked using the Kolmogorov-Smirnov test, followed by the Mann-Whitney *U* test where appropriate considering the non-normality detected for the scale variables (*P*<.001). Multivariate analysis through linear regression was used to explore factors that showed potential significance in univariate analyses (*P*<.100) with collinearity testing using the Variance Inflation Factor (VIF). A *P*-value of less than .050 was considered statistically significant. Statistical analysis was conducted using IBM SPSS Statistics for Windows, Version 26.0.

## Results

### The prevalence of tobacco use in the study sample and its associated determinants

The final study sample comprised 671 university students, with a mean age of 21.04±1.84 years, a majority of females (n=424, 63.2%), and students at public universities (n=498, 74.2%). Additionally, the study sample comprised a variety of students from different disciplines with the most common being medical students (n=236, 35.2%), followed by students in scientific schools (n=189, 28.2%), students in humanities schools (n=134, 20.0%), and students in health schools other than medical schools (n=112, 16.7%).

Moreover, the majority of participants reported parental education at the undergraduate level (for father education, 412 (61.4%) were undergraduates and for mother education 409 (61.0%) were undergraduates). A majority of the study sample reported a middle monthly income for the family (n=510, 76.0%) and a middle level of monthly monetary allowance (n=479, 71.4%). Slightly more than a third of the study participants had a family member (parents or siblings) who work as health care workers (n=226, 33.7%). A majority of the study participants reported having a family member (parents or siblings) who currently consume tobacco (n=484, 72.1%).

In the study sample, the overall prevalence of tobacco use of any kind was 30.0% (95% confidence interval (CI): 26.6%–33.5%), while the prevalence of (e-cigarette use) vaping among those who used any form of tobacco was 62.7% (95% CI: 55.9%–69.2%).

For tobacco use of any kind, univariate analysis showed that the following variables were associated with a higher tobacco use: (1) being a male compared to females (44.9% vs. 21.2%, *P*<.001); (2) enrollment in a private university compared to public universities (39.9% vs. 26.5%, *P*<.001); (3) studying at a humanities school compared to scientific schools, health schools other than medicine, and medical schools (41.0% vs. 37.0% vs. 26.8% vs. 19.5%, *P*<.001); (4) having a family member who currently smokes (35.1% vs. 16.6%, *P*<.001); (5) having a high self-reported monthly monetary allowance compared to low and middle allowances (41.6% vs. 35.0% vs. 26.7%, *P*=.009); (6) having a high self-reported monthly family income compared to low and middle income (40.0% vs. 35.3% vs. 27.3%, *P*=.021); and older age (mean: 21.3 years vs. 20.9 years, *P*=.043, **Table 1**).

**Table 1.** Association between study variables and the consumption of any kind of tobacco products and with vaping.

| Variable | Category | Currently, do you smoke any kind (cigarettes, hookah, e-cigarettes)? |  | *P* value | Are you currently using e-cigarettes (vaping)? |  | *P* value<sup>b</sup> |
| --- | --- | --- | --- | --- | --- | --- | --- |
|  |  | Yes | No |  | Yes | No |  |
|  |  | Count (%) | Count (%) |  | Count (%) | Count (%) |  |
| Age | Mean±SD<sup>a</sup> | 21.3±1.97 | 20.9±1.77 | .043 | 21.3±2.04 | 21.3±1.86 | .881 |
| Sex | Male | 111 (44.9) | 136 (55.1) | <.001 | 75 (67.6) | 36 (32.4) | .112 |
|  | Female | 90 (21.2) | 334 (78.8) |  | 51 (56.7) | 39 (43.3) |  |
| University | Public | 132 (26.5) | 366 (73.5) | <.001 | 83 (62.9) | 49 (37.1) | .938 |
|  | Private | 69 (39.9) | 104 (60.1) |  | 43 (62.3) | 26 (37.7) |  |
| School/Faculty | Medicine | 46 (19.5) | 190 (80.5) | <.001 | 26 (56.5) | 20 (43.5) | .048 |
|  | Health (non-medical) | 30 (26.8) | 82 (73.2) |  | 16 (53.3) | 14 (46.7) |  |
|  | Scientific | 70 (37.0) | 119 (63.0) |  | 53 (75.7) | 17 (24.3) |  |
|  | Humanities | 55 (41.0) | 79 (59.0) |  | 31 (56.4) | 24 (43.6) |  |
| Father education | High school or less | 45 (31.5) | 98 (68.5) | .382 | 20 (44.4) | 25 (55.6) | .015 |
|  | Undergraduate | 116 (28.2) | 296 (71.8) |  | 80 (69.0) | 36 (31.0) |  |
|  | Postgraduate | 40 (34.5) | 76 (65.5) |  | 26 (65.0) | 14 (35.0) |  |
| Mother education | High school or less | 62 (33.3) | 124 (66.7) | .256 | 38 (61.3) | 24 (38.7) | .935 |
|  | Undergraduate | 113 (27.6) | 296 (72.4) |  | 71 (62.8) | 42 (37.2) |  |
|  | Postgraduate | 26 (34.2) | 50 (65.8) |  | 17 (65.4) | 9 (34.6) |  |
| Do you have any family members (parents, siblings) who work in health care? | Yes | 73 (32.3) | 153 (67.7) | .345 | 46 (63.0) | 27 (37.0) | .942 |
|  | No | 128 (28.8) | 317 (71.2) |  | 80 (62.5) | 48 (37.5) |  |
| How would you generally describe your family's monthly income? | Low | 18 (35.3) | 33 (64.7) | .021 | 9 (50.0) | 9 (50.0) | .483 |
|  | Middle | 139 (27.3) | 371 (72.7) |  | 88 (63.3) | 51 (36.7) |  |
|  | High | 44 (40.0) | 66 (60.0) |  | 29 (65.9) | 15 (34.1) |  |
| How would you generally describe your monthly monetary allowance? | Low | 36 (35.0) | 67 (65.0) | .009 | 20 (55.6) | 16 (44.4) | .492 |
|  | Middle | 128 (26.7) | 351 (73.3) |  | 84 (65.6) | 44 (34.4) |  |
|  | High | 37 (41.6) | 52 (58.4) |  | 22 (59.5) | 15 (40.5) |  |
| Do you have any family members (parents, siblings) who currently smoke? | Yes | 170 (35.1) | 314 (64.9) | <.001 | 101 (59.4) | 69 (40.6) | .025 |
|  | No | 31 (16.6) | 156 (83.4) |  | 25 (80.6) | 6 (19.4) |  |

<sup>a</sup> SD: Standard deviation; <sup>b</sup> *P* values were calculated using the Chi square and Mann Whiteny *U* tests.

Univariate analysis also showed that the following variables were associated with higher probability of e-cigarette use (vaping): (1) father education as undergraduate or postgraduate compared to high school or less level of education (69.0% vs. 65.0% vs. 44.4%, *P*=.015); lack of family members (parents or siblings) who currently smoke compared to having a family member who currently smokes (80.6% vs. 59.4%, *P*=.025); and (3) being a student at scientific schools compared to being a medical student, a student in humanities schools, a student in health schools other than medicine (75.7% vs. 56.5% vs. 56.4% vs. 53.3%, *P*=.048, **Table 1**).

### Psychometric properties of the novel VAPeS instrument

The psychometric evaluation of the novel survey instrument used to assess attitudes and perceptions related to vaping among university students (VAPeS) revealed robust results as indicated by the Kaiser-Meyer-Olkin Measure of Sampling Adequacy which was high at 0.823 suggesting that the sample size was adequate for the analysis performed. Bartlett’s Test of Sphericity was significant (χ^2^=1539.159, *P*<.001), which confirmed that the correlation matrix was not an identity matrix and was suitable for factor analysis.

To extract factors from the survey items PCA was employed, with a total of 23 items loading significantly on five components with Eigenvalues >1.0, with five-factor solution explaining a cumulative total of 64.51% of the variance in responses. The first component accounted for 31.79% of the variance, the second 12.81%, the third 7.63%, the fourth 6.38%, and the fifth 5.90% (**Fig 1**).

**Fig 1.**
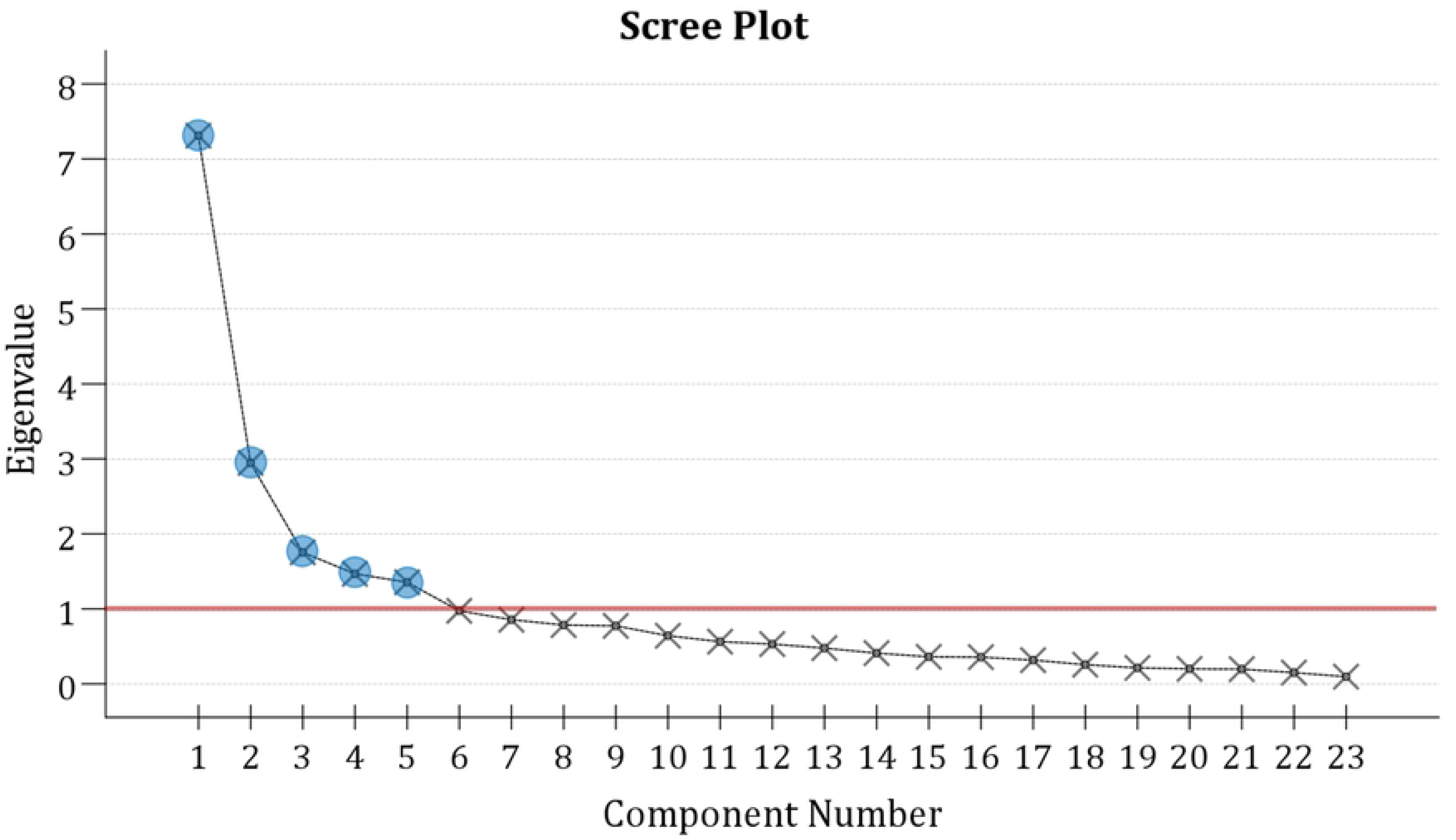
Scree plot of the Vaping Attitude and Perceptions Scale (VAPeS) 23 items using principal component analysis.

The pattern matrix revealed clear loadings of items on respective factors which is suggestive of an acceptable construct validity. The first construct comprising 8 items with factors loading in the range of 0.889–0.557 showed a Cronbach’s α value of 0.888. This construct was termed “Social Influence” based on examination of the items of the construct which were: (1) My vaping experience is influenced by the behavior in my family; (2) I feel pressured to smoke by my peers; (3) My vaping experience is influenced by the behavior of my friends; (4) My role models in life consume tobacco in any form; (5) Vaping is socially accepted among my peers; (6) The portrayal of vaping in movies and series influence how I perceive it; (7) The social media portrayal influence how I perceive vaping; and (8) I believe that vaping helps in social acceptance.

The second construct comprising three items was termed “Harm Perception” with factor loading range of 0.898–0.577, and a Cronbach’s α=0.756: (1) I believe that vaping is less harmful compared to cigarettes; (2) I believe that vaping is less harmful compared to Narghile; and (3) E-cigarettes (vaping) can help smokers quit smoking or cut down on smoking. The third construct comprising two items with factor loadings of 0.638 and 0.587 and Cronbach’s α=0.688 was termed “Vaping Pleasure”: (1) I find vaping enjoyable; and (2) I find vaping satisfying.

The fourth construct comprising six items with factor loadings range of 0.528–0.808, with a Cronbach’s α=0.673 was termed “Behavioral Influences”: (1) I believe that vaping is associated with health risks; (2) I believe that vaping can lead to nicotine addiction; (3) I am aware of anti-smoking policies within my university; (4) Academic stress affects my vaping habits; (5) I believe that vaping helps in stress reduction; and (6) I can easily obtain e-cigarettes.

Finally, the fifth construct comprising four items with factor loadings range of 0.859–0.668 showed a Cronbach’s α value of 0.792 and was termed “Economic and Self-Efficacy Factors”: (1) I believe that vaping is less costly compared to cigarettes; (2) I believe that vaping is less costly compared to Narghile; (3) I feel confident in my ability to resist vaping in social situations; and (4) I feel confident in my ability to quit vaping altogether.

### The determinants of attitude to vaping using the VAPeS scale in univariate analysis

In univariate analysis, the following factors were significant determinants of agreement with the VAPeS items. Male sex (53.3% vs. 31.4% in females, *P*=.015); and higher scores on the following VAPeS constructs: (1) Harm Perception (mean: 3.96±0.82 for the agreement vs. 3.06±0.83 for the disagreement/neutral, *P*<.001); (2) Vaping Pleasure (mean: 4.18±0.85 for the agreement vs. 3.44±0.82 for the disagreement, *P*<.001); (3) Behavioral Influence (mean: 4.52±0.42 for the agreement vs. 3.84±0.75 for the disagreement, *P*<.001); (4) Economic Factors and Self-Efficacy (mean: 4.24±0.79 for the agreement vs. 3.41±0.85 for the disagreement, *P*<.001); and (5) Social Influence (mean: 3.95±0.78 for the agreement vs. 2.62±0.65 for the disagreement, *P*<.001, **Table 2**).

**Table 2.** Association between various study variables with agreement with overall VAPeS categories.

| Categorical variable | Category | VAPeS <sup>a</sup> category |  | *P* value |
| --- | --- | --- | --- | --- |
|  |  | Disagreement/neutral<br>(1–3.67) | Agreement<br>(3.68–5) |  |
|  |  | Count (%) | Count (%) |  |
| Sex | Male | 35 (46.7) | 40 (53.3) | .015 |
|  | Female | 35 (68.6) | 16 (31.4) |  |
| University | Public | 48 (57.8) | 35 (42.2) | .475 |
|  | Private | 22 (51.2) | 21 (48.8) |  |
| School/Faculty | Medicine | 17 (65.4) | 9 (34.6) | .494 |
|  | Health (non-medical) | 9 (56.3) | 7 (43.8) |  |
|  | Scientific | 30 (56.6) | 23 (43.4) |  |
|  | Humanities | 14 (45.2) | 17 (54.8) |  |
| Father education | High school or less | 13 (65.0) | 7 (35.0) | .427 |
|  | Undergraduate | 41 (51.2) | 39 (48.8) |  |
|  | Postgraduate | 16 (61.5) | 10 (38.5) |  |
| Mother education | High school or less | 20 (52.6) | 18 (47.4) | .854 |
|  | Undergraduate | 41 (57.7) | 30 (42.3) |  |
|  | Postgraduate | 9 (52.9) | 8 (47.1) |  |
| Do you have any family members (parents, siblings) who work in healthcare? | Yes | 26 (56.5) | 20 (43.5) | .869 |
|  | No | 44 (55.0) | 36 (45.0) |  |
| How would you generally describe your family's monthly income? | Low | 7 (77.8) | 2 (22.2) | .298 |
|  | Middle | 49 (55.7) | 39 (44.3) |  |
|  | High | 14 (48.3) | 15 (51.7) |  |
| How would you generally describe your monthly monetary allowance? | Low | 8 (40.0) | 12 (60.0) | .055 |
|  | Middle | 53 (63.1) | 31 (36.9) |  |
|  | High | 9 (40.9) | 13 (59.1) |  |
| Do you have any family members (parents, siblings) who currently smoke? | Yes | 55 (54.5) | 46 (45.5) | .617 |
|  | No | 15 (60.0) | 10 (40.0) |  |
| **Scale variable** |  | **Mean±SD <sup>b</sup>** | **Mean±SD** | ***P* value** |
| Age |  | 21.56±2.3 | 21.02±1.63 | .218 |
| Harm Perception |  | 3.06±0.83 | 3.96±0.82 | <.001 |
| Vaping Pleasure |  | 3.44±0.82 | 4.18±0.85 | <.001 |
| Behavioral Influence |  | 3.84±0.75 | 4.52±0.42 | <.001 |
| Economic Factors and Self-Efficacy |  | 3.41±0.85 | 4.24±0.79 | <.001 |
| Social Influence |  | 2.62±0.65 | 3.95±0.78 | <.001 |

<sup>a</sup> VAPeS: Vaping Attitude and Perceptions Scale; <sup>b</sup> SD: Standard deviation; <sup>c</sup> *P* values were calculated using the Chi square and Mann Whiteny *U* tests.

### The determinants of attitude to vaping using the VAPeS scale in multivariate analysis

The linear regression analysis to assess the determinants of VAPeS categories among the participating students who indicated the current practice of vaping, yielded a significant model as indicated by an F-test value of 24.029 (*P*<.001), and the model explained 58.8% of the variance (R²=0.588, adjusted R²=0.563). The model indicated that the strongest predictors of attitude to vaping were “Social Influence” (β=0.494, *P*<.001), “Behavioral Influence” (β=0.206, *P*=.008), and sex which had a modest but significant negative influence (β=−0.154, *P*=.018, **Table 3**).

**Table 3.**
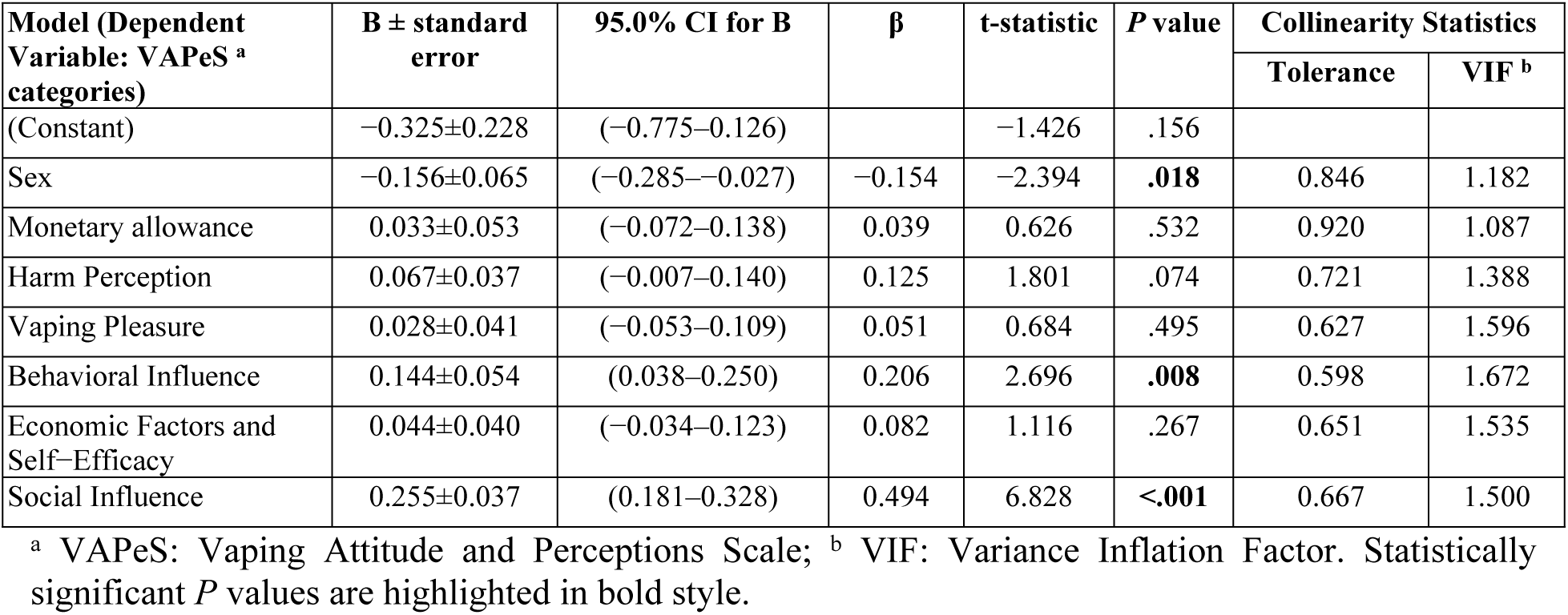
Multivariate analysis for the determinants of vaping in the study sample based on regression analysis.

## Discussion

The principal objective of this study was to develop and validate a comprehensive survey instrument based on the TPB to analyze vaping behaviors among university students in Jordan. This objective is of particular relevance in Jordan among other Arab countries given the evolving patterns of tobacco use in the Middle East countries, where vaping has emerged as a prevalent alternative to traditional methods of tobacco consumption [8,31,37,42]. The selection of university students as the target study population was based on the recognition of this demographic as a susceptible group to trends in vaping due to its accessibility, perceived social acceptability, and aggressive marketing campaigns through channels commonly engaged by younger individuals, such as social media as elaborated by Struik et al. [43].

The findings from our study revealed a 30.0% prevalence of tobacco use among university students in Jordan, with 62.7% of those users reporting current use of vaping. In contextualizing our findings within the global setting, a recent systematic review by Salari et al. provided a comparative benchmark, revealing a global prevalence of vaping use among younger individuals at 16.8% for ever vaping users and 4.8% for current vaping users [44]. This highlights a significant engagement with vaping among youth worldwide, albeit lower than the prevalence observed in our Jordanian university students cohort which was estimated at 18.8% current vaping users. Furthermore, the systematic review by Salari et al. aligned with our observations of sex differences in vaping behaviors, with higher prevalence rates of vaping among males compared to females [44]. Specifically, males showed vaping prevalence rates of 18.8% for ever vaping users and 4.9% for current vaping users compared to 27.4% of current vaping male users in our study [44]. In contrast, young females had significantly lower rates at 9.9% for ever users and 1.6% for current users compared to 12.0% of current vaping female users in our study [44]. These findings highlight the sex-specific dynamics of vaping, suggesting that interventions might need to be tailored differently for young males and females to effectively address these disparities in vaping behavior.

In line with our current findings, previous research within Jordan also highlighted the high prevalence of vaping with significant sex disparities in vaping behaviors. Specifically, a study by Al-Balas et al. reported an 18% prevalence of vaping nationally, with a notably higher prevalence among males (24.9%) compared to females (3.5%) [45]. Moreover, approximately 47.3% of Jordanian participants perceived vaping as less detrimental than traditional cigarette smoking which might explain the upward trend in adopting vaping in the country [45].

Additionally, a recent narrative review focusing on vaping trends across Arab countries further supports the notion of a rising vaping prevalence in the region, which was noted to exceed 25% among the population in studies conducted between 2019 and 2021 [38]. Such trends support the finding of our study and highlight the critical need for targeted public health interventions and ongoing research to address the complex cultural and behavioral changes in tobacco use patterns in the Middle East with the rise of vaping [37].

In this study, the use of a novel survey instrument based on the TPB enabled a comprehensive assessment of the various psychological and social factors that could influence vaping attitudes and behavior. Based on this theoretical framework, we unraveled five factors that would drive the attitude to vaping among university students, namely, Harm Perception, Vaping Pleasure, Behavioral Influence, Economic Factors and Self-Efficacy, and Social Influence. This result appears fathomable considering the diverse motivations that could influence vaping, which range from sensory pleasure to economic considerations. Specifically, Harm Perception and Vaping Pleasure directly address the cognitive considerations and emotional rewards associated with vaping, which are crucial in forming individual attitudes to vaping. Recent studies highlighted a growing awareness of the potential harms of vaping, which would further influence these attitudes [46,47]. Behavioral Influence and Social Influence highlight the significant impact of environmental (e.g., stress) and social contexts, suggesting that vaping behaviors are not only personal choices but also social phenomena which is consistent with the results of a study by Piombo et al. highlighting the role of social factors in vaping initiation [48]. Additionally, Valente et al. highlighted the influence of social networks on e-cigarette use among the youth, which should be considered in public health prevention and cessation programs [49]. Finally, the inclusion of Economic Factors and Self-Efficacy reflects practical and psychological dimensions of vaping, indicating that both cost-benefit analyses and perceived personal control and curiosity factors could play important roles in the decision-making processes related to vaping as elaborated on by Scheinfeld et al. [17]. The multifactorial model identified in the VAPeS scale could provide a robust framework for understanding the complex interplay of factors that would influence vaping, which is important to design tailored public health interventions to address the increasing trend of vaping [16].

In our study, multivariate analysis identified Social Influence as a crucial determinant of vaping behaviors, significantly shaped by peer behavior, media portrayals, and social acceptance. This factor is particularly important among the youth, who are deeply influenced by social norms and media. Additionally, Behavioral Influences, which include health risk awareness and personal vaping experiences, also significantly affected vaping attitudes and behaviors, consistent with the TPB. These findings highlighted how personal perceptions of risk, even if vaping is seen as less harmful could play a key role in individual vaping decisions which should be considered in the intervention measures.

Finally, it is important to consider the results of this study in light of the following limitations. The use of a convenience sampling strategy, while efficient, may limit the generalizability of the findings. Additionally, selection bias is expected considering the female predominance in the sample which may have resulted from a dominance of female authors. Moreover, the cross-sectional design of the study restricts the ability to infer causality between the identified factors and vaping behavior.

To conclude, this study provided a validated tool for future research on vaping behaviors among young adults. We advocate for the broader adoption of the VAPeS scale to further explore the long-term effects of vaping and its dual role as both a potential smoking cessation aid and a gateway to nicotine addiction. Future research should also assess the psychological and socio-cultural determinants of vaping to enhance the effectiveness of targeted interventions and policy actions to address the rising vaping epidemic.

## Acknowledgments

NA

## Data Availability

The data presented in this study are available on request from the corresponding author (MS).

## Author contribution

**Conceptualization:** Muna Barakat and Malik Sallam

**Data curation:** Muna Barakat, Roa’a Abuarab, Baraah Alkharabsheh, Nada Budair, Mais Fareed, Rahma Kharabsheh, Dana Oshroq Laban, Malik Sallam

**Formal analysis:** Malik Sallam

**Investigation:** Muna Barakat, Roa’a Abuarab, Baraah Alkharabsheh, Nada Budair, Mais Fareed, Rahma Kharabsheh, Dana Oshroq Laban, Malik Sallam

**Methodology:** Muna Barakat, Roa’a Abuarab, Baraah Alkharabsheh, Nada Budair, Mais Fareed, Rahma Kharabsheh, Dana Oshroq Laban, Malik Sallam

**Project administration:** Malik Sallam

**Software:** Malik Sallam

**Resources:** Muna Barakat and Malik Sallam

**Supervision:** Malik Sallam

**Validation:** Muna Barakat and Malik Sallam

**Visualization:** Malik Sallam

**Writing – original draft:** Malik Sallam

**Writing – review & editing:** Muna Barakat, Roa’a Abuarab, Baraah Alkharabsheh, Nada Budair, Mais Fareed, Rahma Kharabsheh, Dana Oshroq Laban, Malik Sallam

